# Therapeutic impact of dance therapy on adult individuals with psychological trauma: a systematic review

**DOI:** 10.1101/2022.10.27.22281614

**Authors:** Crystal Tomaszewski, Rose-Angélique Belot, Aziz Essadek, Héloïse Onumba-Bessonnet, Christophe Clesse

## Abstract

**Context:** Dance therapy is a therapeutic tool adapted for people that struggle with classical language-based therapeutic devices (e.g., people facing migration). As dance therapy significantly mobilizes mental, physiological, and somatic dimensions affected by psychological trauma, its therapeutic effect on psychological trauma needs to be evaluated.

**Objectives:** To identify the potential therapeutic effects of dance therapy in adults with psychological trauma (trauma-focused and non-trauma-focused impact) as well as the barriers and facilitators associated with its therapeutic employment.

**Method:** Articles published between 2000 and 2022 have been selected with the help of five relevant keyword combinations applied on seven databases. Two reviewers independently screened titles and abstracts against inclusion and exclusion criteria. Bias evaluation has been conducted with the help of the NIH and JBI. A report of the results has been organized with the help of thematic analysis.

**Results:** Of the thirteen articles included, only one case study directly reports a diminution of pathognomic symptoms of trauma. Other studies present improvements in key dimensions targeted by non-trauma-focused treatment: bodily sensations and perceptions, psychological processes, and interpersonal skills. These improvements depend on the skill set of the therapists and the stability of the intervention.

**Conclusion:** Dance therapy is an effective non-trauma-focused treatment, as it mobilizes the mandatory dimensions for the management of psychological trauma. It is specially adapted to women survivors of violence, and people facing migration. Researchers and professionals should investigate dance therapy as a trauma-focused treatment.

**Highlights:**

- While the non-trauma-focused effects of dance therapy have been particularly evaluated, there is a lack of evidence about studies evaluating the effect of dance therapy as a trauma-focused intervention.
- Dance therapy improves key dimensions targeted by non-trauma-focused intervention therapies: sensory-motor perceptions, motor skills, identification, expression, and externalization of emotions, reflexivity, creativity, interpersonal skills, and verbal expression.
- The effectiveness of dance therapy interventions is correlated with the therapist’s skill set, the intervention’s stability, and the patient’s flow state.

## Introduction

Since the 1940s, dancing activities have been considered a therapeutic tool in Western societies (Dunphy et al., 2022). Dance therapy interventions are usually implemented in health (psychiatric hospitals), social (rehabilitation centers, drop-in centers), and educational (schools) settings. These interventions are carried out by professionals that commonly work in artistic and therapeutic fields. Dance therapy (DT) encompasses Dance/Movement Therapy (DMT), defined as “the psychotherapeutic use of movement to promote the emotional, social, cognitive, and physical integration of the individual” (American Dance Therapy Association, n.d.). Dance therapy mobilizes the individual’s cognitive sphere (learning ability, attention, memory, rhythmic motor coordination, and visual-spatial ability), as well as the creative and emotional sphere (movement improvisation through music, imagination, emotion, and social interaction) (Pessoa et al., 2019). To date, meta-analyses and randomized clinical trials have evaluated the use of dance therapy and demonstrated its positive impact on physical and mental health (Koch, Riege, et al., 2019; Wang et al., 2022). Studies have shown that dance therapy is an effective way to improve mental and physical health, especially for individuals without psychopathological disorders (children, adults, and the elderly), with psychopathological disorders (adults hospitalized in psychiatry), and with neurocognitive disorders (neuropsychological deficits, physical and psychological disabilities) (Ritter & Low, 1996).

The positive therapeutic effects of dance therapy on physical and mental health make it a relevant psychotherapeutic treatment tool. Because of its emphasis on the body, dance therapy is particularly well-suited to specific clinical settings in which individuals have difficulty using conventional language-based psychotherapeutic approaches (Taylor et al., 2020; Van de Kamp et al., 2019). Consequently, health and social service institutions are increasingly looking to dance therapy to address their patients’ needs. Today, several institutions that implement and explore the effect of dance therapy protocols consider dance therapy as beneficial for individuals with psychological trauma (Levine & Land, 2016; Pierce, 2014). A psychological trauma is a significant psychological shock caused by one or more critical events that can alter an individual’s psychic and physiological functioning (Perrotta, 2020). Psychological trauma thus includes post-traumatic stress disorder (PTSD) and complex post-traumatic stress disorder (CPTSD) (American Psychiatric Association, 2013; World Health Organization, 2019). With an estimated prevalence of 3.9% in the general population, and 5.6% among people who have been exposed to traumatic events in their lives, PTSD affects women twice as much as men (Koenen et al., 2017). It particularly affects people facing migration with a range of PTSD prevalence estimated between 2.2% and 9.3% (Silva et al., 2021). The typical symptoms of PTSD (intrusion, avoidance, disturbances in cognition and mood, changes in arousal and reactivity) are very disabling, especially when mechanisms of traumatic revivification and dissociation phenomena are present (American Psychiatric Association, 2013). Psychological trauma significantly impacts the mental, physiological, and somatic (cardiovascular system, immune status, metabolic systems, and inflammatory responses) dimensions of the subject (American Psychiatric Association, 2013; Hillis et al., 2016; O’Donnell et al., 2021). Because of these multiple consequences, psychological trauma can alter the therapeutic and medical work (Green et al., 2016), given that it hinders communication between the caregiver and the patient (Bassuk et al., 2001) while contributing to the patient’s disengagement (Green et al., 2016).

From a psychotherapeutic point of view, dance therapy mobilizes the cognitive, creative, and sensorial dimensions of the subject, known to be particularly targeted by psychotherapeutic intervention devoted to psychological trauma (Levine & Land, 2016; Pessoa et al., 2019). Psychological trauma can be treated with trauma-focused and non-trauma-focused treatment psychotherapies (Watkins et al., 2018). Non-trauma-focused treatment is recommended when trauma-focused and/or pharmacotherapy are not available or declined by the patient (VA/DOD, 2017). Non-trauma-focused intervention for the management of PTSD focuses on improving multiple key dimensions of the patient (VA/DOD, 2017; Watkins et al., 2018), such as mobilization of the body at a somatic level to address deep-rooted physiological responses to risk, relaxation, diminution of stress, and interpersonal therapy (Grabbe & Miller-Karas, 2018; Watkins et al., 2018). Mobilizing these specific key dimensions are essential aspect of the psychotherapeutic resolution of psychological trauma in the case of non-trauma-focused-treatment.

To promote better therapeutical practices adjusted to the clinical particularities of patients, scientific research then needs to better understand and isolate the potential therapeutical effects of dance therapy on specific populations as well as their most associated mental health difficulties. The primary objective of this article is therefore to identify the potential therapeutic effects of dance therapy on adults with psychological trauma (trauma-focused and non-trauma-focused effects). Its second objective is to identify the barriers and facilitators associated with the impact of dance therapy on adults suffering from psychological trauma. This study aims to guide future studies on the effectiveness of dance therapy in the management of psychological trauma, refine the quality of clinical referrals, and provide theoretical support for clinical dance therapy practices.

## Method

### Search strategy

The present qualitative systematic review protocol, registered in PROSPERO under the reference XXXXXXXXXXXXXX, is a part of the research project VaDDanC (Validation of a Dance-therapy Device in Co-therapy), funded by LOBA association. The aim of the present study is to evaluate the potential therapeutic impact of dance therapy on adults suffering from psychological trauma.

The search strategy has been conducted in a double-blind manner by two authors (CT and CC) (Figure 1). They screened the articles published between 2000 and June 2022 with the help of a list of French and English keywords: dance/danse”, “therapy/therapies, “trauma/trauma”, “intervention/intervention”, “group/groupe”, “violence/violence”. Those keywords have been organized into 5 different keyword combinations applied on 7 databases: Cochrane, Embase, PubMed, Web of Science, PsycINFO, PsycArticles, and ScienceDirect (supp data 1). Complementary research performed on the 20 first result pages of Google Scholar for those keyword combinations, and in the reference list of selected articles, ensured that no additional paper has been missed by the search strategy.

**Figure 1.**
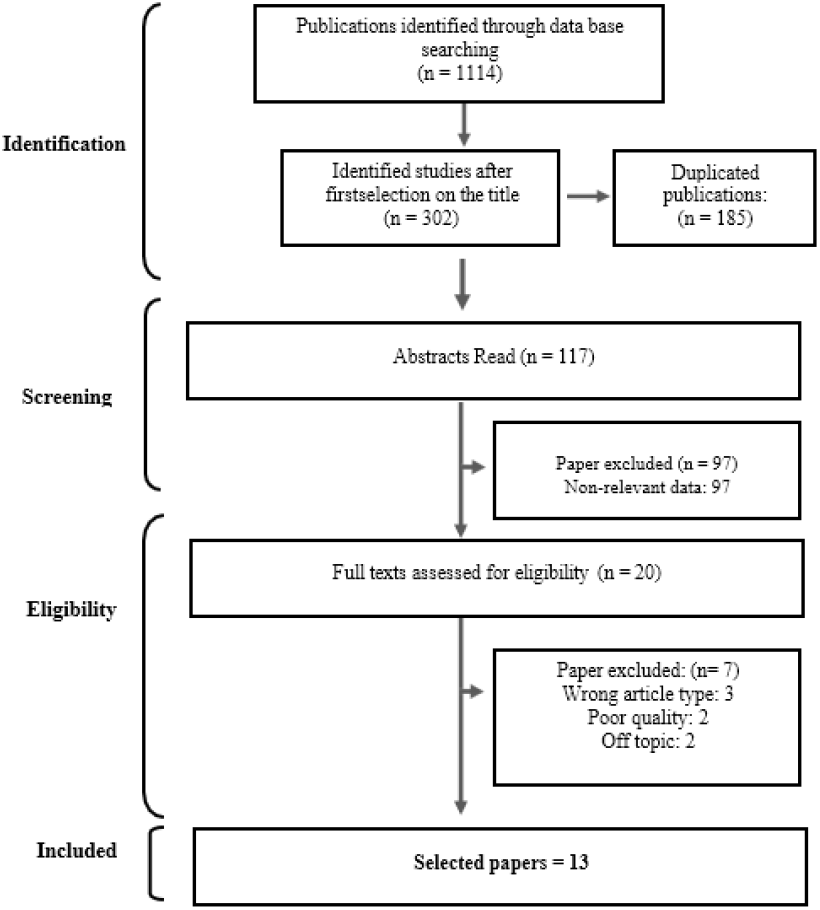
Flow diagram of the selection process.

### Inclusion criteria and bias assessment

The inclusion criteria of the study are: the scope of the study (over 18 adult-focused studies, individuals with psychological traumatism, dance therapy, and dance movement therapy) and articles published between 2000 and June 2022. The exclusion criteria are: studies focusing on children and adolescents under 18, out-of-scope articles, literature reviews, book chapters, books, published theses, dissertations, conference abstracts, guides, and expert reports. Bias assessment (Table 1) has been carried out by two researchers (XX and XX) with the help of two assessment tools: the NIH (National Institutes of Health, 2014) and the JBI (Aromataris & Munn, 2020). Both tools were used when permitted by the article design, and the results were cross-tabulated. Case studies have been assessed with the JBI only, as the NIH is not designed for this purpose.

**Table 1:**
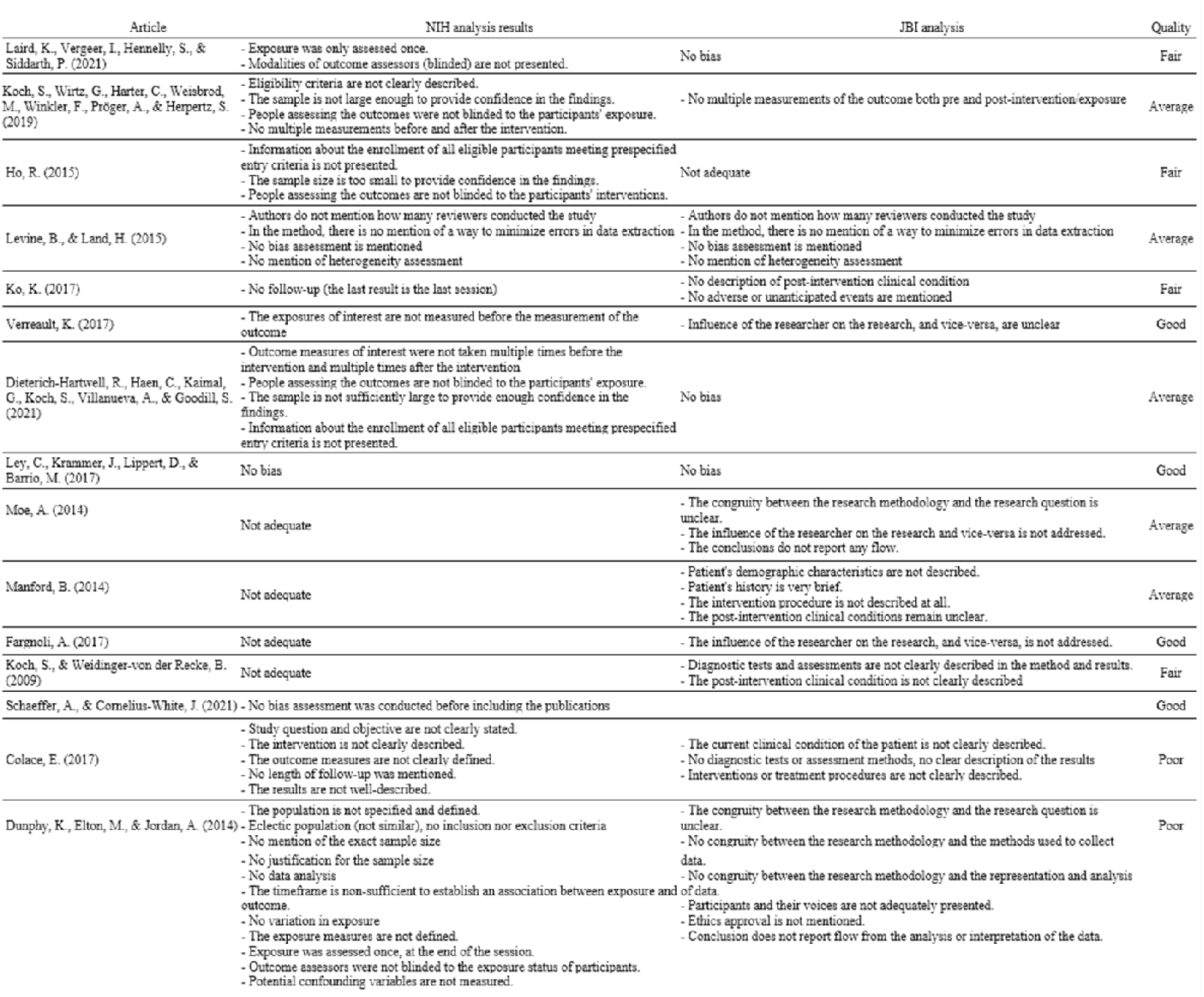
Bias analysis.

At the end of the selection process, authors extracted the following available data (Table 2 & Table 3): authors, date of publication, the title of the publication, country undertaking the study, objectives, hypotheses, type of article, population, study parameters, type of analysis, main results (Table 2), secondary results, and authors’ main interpretations. At last, a thematic analysis (Braun & Clarke, 2006) based on the extracted data isolated relevant sub-themes and themes exploring the impact, associated factors, and barriers associated with dance therapy and psychological trauma.

**Table 2:**
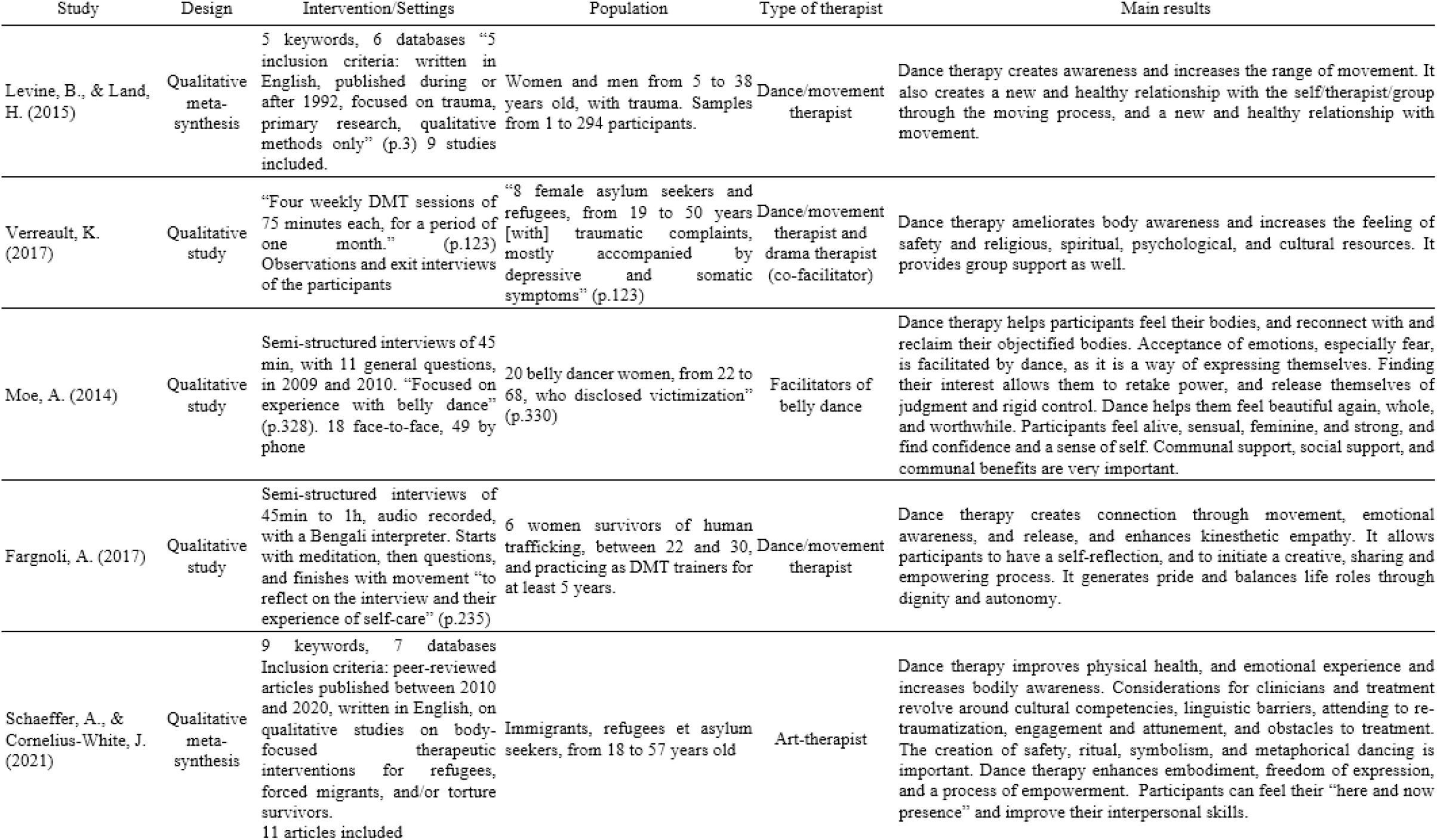
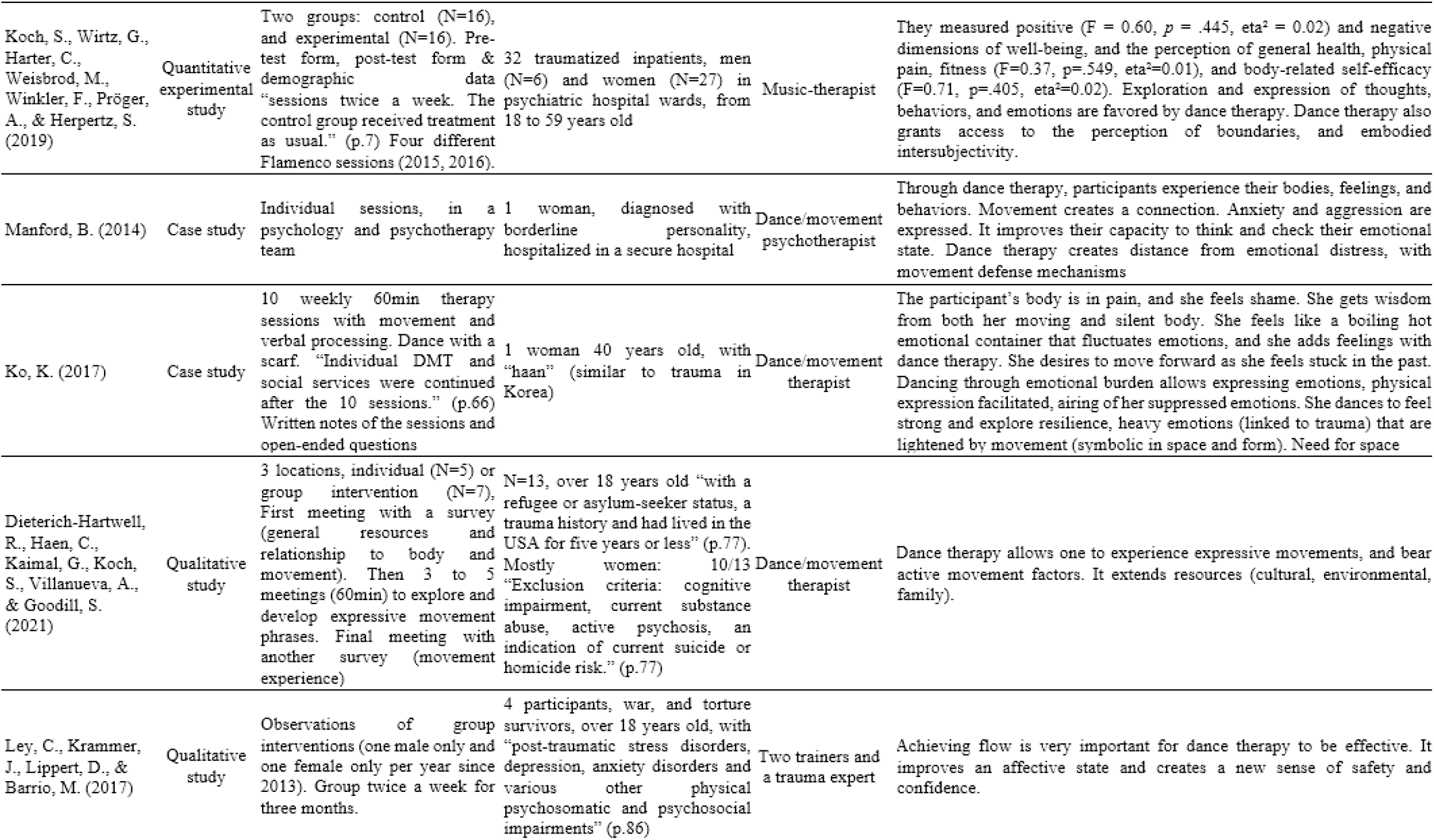
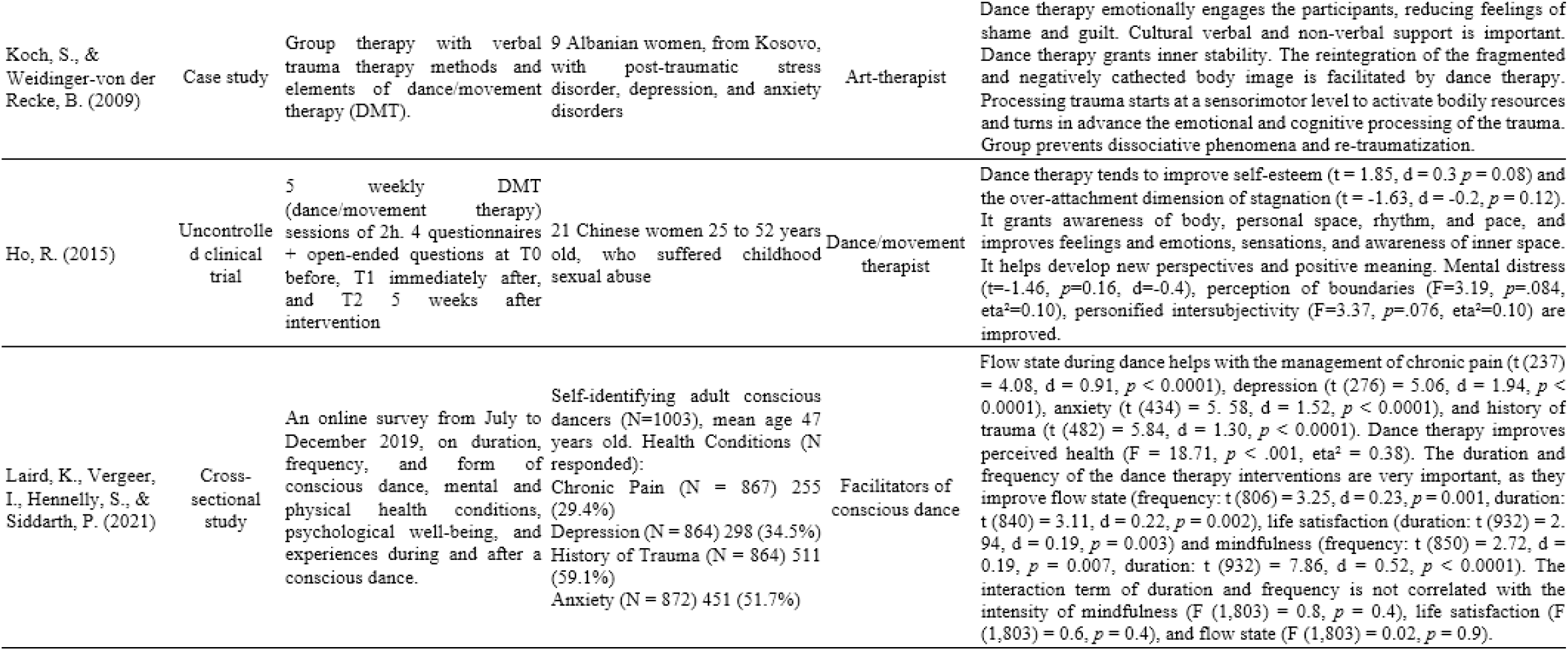
Selected articles

**Table 3:** Results of the thematic analysis

| Themes | Sub-themes | Frequency |
| --- | --- | --- |
| Body and space | Bodily sensations and perceptions | 9 |
|  | Movement | 5 |
| Individual psychological factors | Emotions | 7 |
|  | Psychopathological symptoms | 3 |
|  | Identity and mental health | 7 |
| Relationship | Relationship with others | 8 |
|  | Self-expression | 3 |
| Group intervention | Security | 4 |
|  | Culture | 3 |
|  | Group characteristics | 4 |
| General Health | Well-being | 4 |
|  | Physical pain | 3 |
| Psychological process | Linking affects and representations | 2 |
|  | Mental movement | 2 |

### Study selection

The database search let the authors screen 1114 articles (supp data 1). Among the 117 selected titles, a dual evaluation using the Rayyan QCRI software (Ouzzani et al., 2016) has been performed on the abstracts. The application of inclusion and exclusion criteria then led to an exclusion of 102 articles. An analysis of bias has been carried out on the 15 remaining articles and led to the exclusion of 2 articles that presented too much methodological bias (the description of the intervention was too vague, the variables were not established, the data collection and analysis did not correspond to the protocol presented, the sample was not described). As a result, 13 articles have been finally included in this qualitative systematic review.

### Data analysis

As data extraction did not isolate enough quantitative data per studied variables (two or more quantitative results per the same variable (Deeks et al., 2022)), a meta-analysis could not be performed. Two researchers conducted a thematic analysis and identified 14 common subthemes grouped into 6 main themes. The results of this qualitative systematic review are then presented and discussed according to the SWiM protocol (Campbell et al., 2020). A list of recommendations for research and clinical practice is also formulated to shed light on future theoretical, psychotherapeutic, and psychopathological directions for dance therapy.

## Results

A total of 13 articles is included in this qualitative systematic review. These studies have been conducted in Europe (Germany/3, The Netherlands/2, Austria/2, The United Kingdom/2), the United States of America (5), and China (1). Selected studies encompass two meta-syntheses (with one including two articles from our selection process (Ley et al., 2017; Verreault, 2017)), 3 quantitative studies (1 monocentric and 2 multicentric), 5 qualitative studies (3 monocentric and 2 multicentric), and 3 monocentric case studies.

Sample sizes of studies range from 1 to 864 including individuals with psychological trauma. The samples are predominantly female (6 papers with exclusively female populations and 7 papers with predominantly female populations). The diagnosis of psychological trauma (PTSD and psychological trauma) has been systematically applied to all study participants and was associated with migration (4 studies), violence in daily life (4 studies), and war survival (2 studies). Data were collected through semi-structured interviews (5 studies), questionnaires (4 studies), and observations (5 studies). The thematic analysis (Braun & Clarke, 2006) isolated 6 major themes: Body and Space, Individual Psychological Factors, Relationship, Group intervention, General Health, and Psychological Processes (Table 3).

### Psychological trauma

Of the 13 selected articles, only one case study (Koch & Weidinger-von der Recke, 2009) directly reports a diminution of traumatic revivification and dissociation after a patient got enrolled in a dance therapy program. This diminution is due to addressing psychological trauma on a bodily level through movement and dance, which helps the patients to regain a positive relationship with their body (ibid). The authors consider that combining verbal and non/verbal dimensions of psychotherapy is essential for the symptom’s diminution, as well as the reconstruction of the fragmented image of the body and the reinforcement of the individual’s resources.

No included study measured other typical symptoms such as intrusion, avoidance, disturbances in cognition and mood, and disabling changes in arousal and reactivity. Despite a lack of evidence directly focusing on symptoms of PTSD and CPTSD, selected studies provided an evaluation of the physical, psychological, and social dimensions affected by psychological trauma. Those key dimensions (relaxation, diminution of stress, interpersonal therapy, mobilization of the body) are targeted by non-trauma-focused treatment because they participate in and facilitate the remission of psychological trauma while being an asset in the recovery process of the subject.

### Body and space

#### Bodily sensations and perceptions

Selected studies demonstrate that dance therapy improves the sensory-motor perceptions of individuals suffering from psychological trauma. Qualitative studies point to improved body awareness and the emergence of a mind-body connection through the sensory experience of movement, rhythm, and relaxation (Fargnoli, 2017; Levine & Land, 2016; Moe, 2014; Schaeffer & Cornelius-White, 2021; Verreault, 2017). Unfortunately, these results are not confirmed by an experimental quantitative study, that evaluated the relation between body self-efficacy and the perceived level of fitness of participants after one Flamenco session (Koch, Wirtz, et al., 2019). The differences between the control group and the Flamenco group were not significant (fitness: F=0.37, p=.549, eta^2^=0.01, and body-related self-efficacy: F=0.71, p=.405, eta^2^=0.02). Two major limitations of this study should yet be reported: the small sample size and the fact that there was only one Flamenco dance session. On another level, the presence of mechanisms associated with bodily experience and the mind-body connection has also been described by a case study article (Manford, 2014) focusing on a patient suffering from a borderline personality disorder. The study highlighted that improvement of the sensory-motor perception could occur even in the case of a severe psychiatric diagnosis.

#### Movement

Included studies demonstrate that dance therapy improves the motor skills of individuals with psychological trauma. The qualitative studies point to an increase in the range of motion, an expansion of movement repertoire, and the creation of a mind-body connection through movement (Dieterich-Hartwell et al., 2021; Fargnoli, 2017; Levine & Land, 2016). Repetition of the sessions is essential for participants to become aware of these improvements (Dieterich-Hartwell et al., 2021). Experience of the moving and silent body has been similarly reported in a case study of a migration-affected woman (Ko, 2017). However, it appears that expression through movement can only occur if the therapist is able to observe and decipher the movement (Fargnoli, 2017; Levine & Land, 2016).

### Individual psychological factors

#### Emotions

When applied to individuals suffering from psychological trauma, dance therapy is associated with an improvement in the identification, expression, and externalization of emotions. The qualitative studies isolated an improvement in an emotional state, a better recognition of emotions, a better focus on emotions, an improvement in emotional release, and the development of kinesthetic empathy (to feel in one’s own body the movement of the other) (Fargnoli, 2017; Ley et al., 2017; Moe, 2014; Schaeffer & Cornelius-White, 2021). Two case studies also noticed that the first emotional expressions after dance therapy involve the expression of fear, shame, and guilt (Ko, 2017; Koch & Weidinger-von der Recke, 2009). Dance therapy also promotes the emergence of psychic processes such as awareness and verbalization of effects, as well as an alleviation of suffering (Ho, 2015; Koch & Weidinger-von der Recke, 2009). The emotional expression promoted by dance therapy also facilitates the processing of the trauma (Ko, 2017) and the improvement of the emotional experience (Schaeffer & Cornelius-White, 2021). However, these improvements are observed during the treatment or just after the last session. The lack of longitudinal observations in the selected studies prevents any conclusion about the persistence of these improvements.

#### Identity and mental health

Our analysis shows that when employed with individuals suffering from psychological trauma, dance therapy mobilizes identity factors and improves their mental health. Qualitative studies indicate that dance therapy mobilizes psychological resources and improves reflexivity and creativity (Fargnoli, 2017; Moe, 2014; Schaeffer & Cornelius-White, 2021; Verreault, 2017). DT also generates a renewed feeling of being alive, a better enjoyment in “the here and now” as well as a facilitated initiation of the process of empowerment. Additionally, dance therapy facilitates the rediscovery of self-encouraged individual and social dimensions (femininity, sensuality, and spirituality) (Moe, 2014; Verreault, 2017). These movements contribute to the reconstruction of the self and increase self-confidence and pride in being oneself. On this point, case studies also show the emergence of a will to move toward the future, an improvement in the ability to think and to be attentive to one’s emotional state (Dieterich-Hartwell et al., 2021; Ko, 2017; Manford, 2014). Those improvements have been assessed as a way to better construct a new internal and psychological stability while helping to construct a new internal and psychological stability (Koch & Weidinger-von der Recke, 2009). In sum, these results converge sufficiently to show a positive impact of dance therapy on the mental health of individuals with psychological trauma.

#### Psychopathological symptoms

Several articles provided an evaluation of specific symptoms of mental health afflictions associated with psychological trauma. A non-randomized clinical trial involving 21 women participating in a dance therapy program demonstrated a non-significant decrease in mental distress before and after participation in a DMT program (t=-1.46, *p*=0.16, d=-0.4) (Ho, 2015). Similar mental health improvements have been also commented on by a case study focusing on a woman diagnosed with borderline personality disorder (Manford, 2014). In parallel, a cross-sectional study (N=1003) associated the improvement in the psychological state with the flow state of participants, defined as a “state in which people are so involved in an activity that nothing else seems to matter; the experience itself is so enjoyable that people will do it even at great cost, for the sheer sake of doing it” (Csikszentmihalyi, 1990) (Laird et al., 2021). In this study, 899 people answered that they had one or more mental health afflictions: traumatic experience (N=511), anxiety (N=451), depression (N=298), chronic pain (N=255), and drug addiction (N=205). Most of the subjects in this study reported that dance helped them to manage their mental health affliction (depression 96.3%, anxiety 96.2%, trauma 94.9%, chronic pain 89.4%, and substance abuse, 87.8%). Flow state has been isolated as a significant facilitator for mental health improvements. The correlation between each mental health affliction variable and flow state was significant for 276 individuals in the management of depression (t (276) = 5.06, d = 1.94, *p* < 0.0001), for 434 in the management of anxiety (t (434) = 5. 58, d = 1.52, *p* < 0.0001), for 482 in managing traumatic experience (t (482) = 5.84, d = 1.30, *p* < 0.0001), and for 237 in managing chronic pain (t (237) = 4.08, d = 0.91, *p* < 0.0001). This study is based on a self-report method that could induce bias in the identification of symptoms (Laird et al., 2021). However, dance therapy improves the management of the symptoms of depression, anxiety, traumatic experience, and chronic pain, as the participants can enter a flow state.

### Relationship

#### Relationship with others

One of the leading themes of the selected article is the evaluation of the impact of dance therapy on interpersonal dimensions and relationships. First, in a non-randomized clinical trial comparing twenty-five women that participated in a Flamenco session vs a control group, a significant improvement (p. <0.10) in their perception of boundaries (F=3.19, *p*=.084, eta^2^=0.10) and personified intersubjectivity (F=3.37, *p*=.076, eta^2^=0.10) has been observed (Koch, Wirtz, et al., 2019). Four qualitative studies contribute to confirming this aspect by showing that dance therapy improves the construction of spatial and bodily limits, and the ability to create links with others (Fargnoli, 2017; Moe, 2014; Schaeffer & Cornelius-White, 2021; Verreault, 2017). These effects are facilitated by the quality of group support (Verreault, 2017), and the attention to the cultural and social characteristics of the participants (Fargnoli, 2017; Moe, 2014; Schaeffer & Cornelius-White, 2021). As demonstrated by a group case study, the presence of peers during the trauma narrative is essential (Koch & Weidinger-von der Recke, 2009). It favors sustained attention and the mirroring of the individual’s emotional state (ibid). In addition, the group setting offers a verbal and/or non-verbal holding function to the individual (ibid). This helps to prevent too much internal rigidity and reduces the phenomenon of dissociation. Dance therapy also encourages the expression of the patient’s need for space (Ko, 2017).

#### Self-expression

Self-expression through dance therapy has been also investigated. In two qualitative studies and a case study, participants with psychological trauma experienced a long-lasting improvement in verbal expression, self-expression, and freedom of speech (Manford, 2014; Moe, 2014; Schaeffer & Cornelius-White, 2021). These improvements are facilitated by a feeling of confidence and safety (ibid).

### Group intervention

#### Security

When combined with the development of the ritual and symbolic aspects of dance, the sense of security experienced in the group contributes to the development of a generalized individual sense of safety (Ley et al., 2017; Schaeffer & Cornelius-White, 2021; Verreault, 2017). This sense of safety then allows to individual resources to emerge in participants (Verreault, 2017) while facilitating the transition through a flow state (Schaeffer & Cornelius-White, 2021). The emergence of these associated factors is facilitated by the permanence and the framework of the interventions (quality of the premises, geographical location, and regularity) (Schaeffer & Cornelius-White, 2021).

#### Culture

As reported by the included studies, cultural, religious, and spiritual support facilitate the management of psychological trauma. This statement is supported by two qualitative studies that show how this factor expands the subject’s own cultural, family, and environmental resources highlighting how the subjects involved in a DT program expand their cultural, family, and environmental resources (Dieterich-Hartwell et al., 2021; Verreault, 2017). Dance therapy is especially useful for trauma management in a multicultural setting, as it mobilizes non-verbal skills while complementing psychosocial interventions through cultural and family support (Dieterich-Hartwell et al., 2021; Koch & Weidinger-von der Recke, 2009; Verreault, 2017).

#### Group characteristics

The specific settings of group intervention have been also reviewed by scientific literature. Duration (dance practice for more than 5 weeks vs. less than 5 weeks) and frequency (dance practice at least once a week vs. twice a month or less) are positively correlated with flow state (frequency: t (806) = 3.25, d = 0.23, *p* = 0.001, duration: t (840) = 3.11, d = 0.22, *p* = 0.002), life satisfaction (duration: t (932) = 2. 94, d = 0.19, *p* = 0.003) and mindfulness (frequency: t (850) = 2.72, d = 0.19, *p* = 0.007, duration: t (932) = 7.86, d = 0.52, *p* < 0.0001) (Laird et al., 2021). The combination of long duration and high frequency of dance sessions however does not intensify their individual positive effect on the intensity of mindfulness (F (1,803) = 0.8, *p* = 0.4), life satisfaction (F (1,803) = 0.6, *p* = 0.4), and flow state (F (1,803) = 0.02, *p* = 0.9). Settings of the interventions are crucial to create a new healthy relationship with oneself, the group, the therapist, and the movements (Levine & Land, 2016; Schaeffer & Cornelius-White, 2021). The quality of these relationships depends on the ability of the therapist to consider cultural dimensions, difficulties in verbal expression, and the risk of repetition of the trauma (Schaeffer & Cornelius-White, 2021). Moreover, the quality of the relationships engaged by the subject is conditioned by the therapist’s ability to identify obstacles, engage with the group, and adapt to the patients (Schaeffer & Cornelius-White, 2021). Therapists play a critical role in the care and must be aware of their countertransference, their use of space, and the theoretical paradigms associated with their movements (Levine & Land, 2016).

### General Health

#### Well-being

Well-being has been assessed through several different variables in quantitative studies (Ho, 2015; Koch, Wirtz, et al., 2019; Laird et al., 2021). Among the well-being evaluated variables, people that practiced dance for more than 5 weeks presented higher mindfulness scores (t (932) = 7.86, d = 0.52, *p* < 0.0001), higher levels of life satisfaction (t (932) = 2.94, d = 0.19, *p* = 0.003) and better flow states (t (840) = 3.11, d = 0.22, *p* = 0.002) (Laird et al., 2021). In a different sample assessed by this same study (Laird et al., 2021), people who dance at least once a week have also higher mindfulness scores (t (850) = 2.72, d = 0.19, *p* = 0.007) and a better flow (t (806) = 3.25, d = 0.23, *p* = 0.001). Two other qualitative studies (Ley et al., 2017; Schaeffer & Cornelius-White, 2021) corroborate these results. The well-being component assessed through perceived health is also ameliorated by dance therapy (F = 18.71, *p* < .001, eta^2^ = 0.38) (Koch, Wirtz, et al., 2019).

Improvements in well-being through dance therapy have not been reported by a non-randomized clinical trial focusing on self-esteem and stagnation, a concept in Eastern medicine that resembles to depression in western medicine (Ho, 2015). The increase in self-esteem presents nonetheless a tendency to statistical significance (t = 1.85, d = 0.3 *p* = 0.08) that does not persist 5 weeks after the end of the program (t = 1.11, d = 0.2, *p* = 0.28). Stagnation Scale’s (Ng et al., 2006) overattachment dimension also did not report a decrease in stagnation (t = -1.63, d = -0.2, *p* = 0.12) whereas a non-statistically significant tendency seems to emerge 5 weeks after the program (t = -1.81, d = -0.3, *p* = 0.08). A similar trend has been reported by a quantitative experimental study showing that positive well-being dimensions are not significantly higher after dance therapy (F = 0.60, *p* = .445, eta^2^ = 0.02) (Koch, Wirtz, et al., 2019).

#### Physical pain

Mainly assessed through self-report, physical pain has also been investigated. In a large sample study (N=1080), the majority of participants with chronic pain (n=255), 89.4% reported being helped in the management of their pain by dance therapy sessions (Laird et al., 2021). Participants who reported improvement in pain management had a better flow state compared to those who reported not being helped (t (237) = 4.08, d = 0.91, *p* < 0.0001) (Laird et al., 2021). This result is complemented by the study focusing on a flamenco session showing that when performing a dance session, participants present less physical pain (F = 4.25, *p* = .048, eta^2^ = 0.12) (Koch, Wirtz, et al., 2019). Cases study’s results also consider that dance therapy sessions can improve emotional pain and consequently can reduce physical pain (Ko, 2017).

### Psychological processes

#### Links between affects and representations

Selected case studies also reported that dance therapy investigates the link between affects and representations through emotional expression (Ko, 2017). As physical activity allows for the expression of repressed emotions and leads to a feeling of liberation and relief, dance therapy can create a safe distance from emotional distress, often described as an overwhelming feeling (Manford, 2014). Both selected case studies consider these mechanisms to occur through the link made by the subject between his affects and his somatic expression (Ko, 2017; Manford, 2014). They identify the outlet function of movement (Ko, 2017) and establish a correlation between physical movements and psychic movements (Manford, 2014).

#### Mental movement

Practicing dance therapy in a group also allows one to explore sensorimotor perceptions, which initiates the processing of the trauma (Koch & Weidinger-von der Recke, 2009). Identifying and activating the processes at the base of the body’s resources enable the process of emotional and cognitive integration of the trauma to begin. Activation of the sensory-motor pole allows for the reintegration of the fragmented and negatively invested body image (Koch & Weidinger-von der Recke, 2009). In addition, dance therapy enables the patient to gain a sense of strength and to express intense emotions linked to past trauma (Ko, 2017). The reduction of symptoms can be associated with the reintegration of fragmented body image and the reinforcement of psychic resources. Movement can have a symbolic function when facilitating the emergence of a diversity of affects that will be subjected to interpretation (Ko, 2017).

## Discussion

The 13 included studies show that, despite not providing typical quantitative measures about trauma-focused interventions, dance therapy appears effective in case of non-trauma-focused treatment. On one hand, dance therapy can improve the reappropriation of the body, the expression of emotions and, ameliorates the management of psychopathological symptoms in people with psychological trauma. Group dance therapy also favors self-expression and the creation of new relationship with others. On the other hand, the feeling of safety and the ability of participants to be in a flow state are known to facilitate the emergence of these non-trauma-focused improvements. At last, those improvements depend on the skill set of the therapists and the stability of the intervention.

As only one case study directly reported a diminution of traumatic revivifications and dissociations (Koch & Weidinger-von der Recke, 2009), it is impossible to draw conclusions about the potential trauma-focused effects of Dance Therapy. These emerging trauma-focused results (*ibid*) still need to be extensively investigated by qualitative and quantitative research that targets a reduction of PTSD and CPTSD cases, as well as a reduction in psychological trauma symptoms. Conversely, key dimensions targeted in the case of non-trauma-focused interventions have been rather investigated by the 13 selected studies. Among the key traumatic dimensions targeted by non-trauma-focused treatments, our analysis shows that dance therapy improves bodily sensations, perceptions & movements, psychological processes, and interpersonal skills (Dieterich-Hartwell et al., 2021; Fargnoli, 2017; Levine & Land, 2016; Schaeffer & Cornelius-White, 2021; Verreault, 2017). These improvements led to consider that dance therapy is efficient in the resolution of psychological trauma and should be considered as a non-focused trauma treatment intervention. Today, non-focused trauma interventions are recommended when trauma-focused treatments are unavailable/unwanted by the patient or as second-line treatment (VA/DOD, 2017). In that context, future recommendations and guidelines devoted to psychological trauma support could also include DT as a non-trauma-focused relevant intervention particularly adjusted to diverse ethnical and cultural populations such as people affected by migration.

Among the key dimensions positively improved by dance therapy, sensory-motor perceptions and motor skills are particularly emphasized. These results have been highlighted by other studies on dance therapy, especially in the case of mental health afflictions prevention policies and when applied to children with disabilities (Koch, Riege, et al., 2019; May et al., 2021). It is indeed established that repeated physical activity is strongly correlated with the development of motor skills (Hirano et al., 2015). By being a potent reminder of the initial sensory-motor skills acquisition, dance therapy also strengthens sensory-motor perceptions (Todd & Lee, 2015). All of those ameliorations then improve the body schema, which in turn promotes the development of motor skills (Maravita & Iriki, 2004). Positive results on this key dimension are considered a great help for psychological trauma management (Grabbe & Miller-Karas, 2018). This, as it helps the subject to redevelop a deep connection with its body and sensations known to be distanced by dissociation and amnesia (Van der Kolk, 2006). Potential emphasizes by group feedback after dance therapy sessions, working with mirroring, weight, rhythm, body perception exercises, and spatial reference can be apply to reinforce these effects (Koch, 2020).

The effects of dance therapy are also mainly dependent on the improvements in the identification, expression, and externalization of emotions. This is consistent with the literature on the effectiveness of dance therapy in the general population (Koch, Riege, et al., 2019; May et al., 2021; Schwender et al., 2018). As dance is a non-verbal medium, it instigates the expression of emotions through the body while initiating the process of emotional identification (San-Juan-Ferrer & Hípola, 2020). Emotional improvements fostered by DT are particularly relevant in the case of traumatic dissociation as this pathognomonic symptom is characterized by a disruption in the integration of emotions (American Psychiatric Association, 2013). Dance therapy can then strengthen the psycho-affective state of the subject (Schaeffer & Cornelius-White, 2021). It can therefore improve the subject’s mentalization abilities frozen by sideration and, enable a reduction of the feelings of shame and guilt often expressed by individuals suffering from psychological trauma (Pugh et al., 2015; Wilson et al., 2006). Clinicians and dance therapists also should expect that the first emotional register presented can be feeling of guilt and shame (Ko, 2017). Similarly, authentic movement exercises with closed eyes, group touching, and without music should be avoided in the early stages of treatment to avoid a risk of retraumatization (Koch, 2020).

A special focus is needed on the ameliorations brought by DT in terms of personal safety, cultural support, and the creation of new interpersonal relationships. These improvements allow for the intrusive re-emergence of the traumatic image, linked to the traumatic memory, to be contained and processed in the group (Kruse et al., 2009). Aside from the implications of these key factors in the management and the resolution of psychological trauma (Watkins et al., 2018), it is possible that holding and containment mechanisms are key facilitators of their apparition, as theorized by Winnicott, and Bion (Bion, 1961; Karampoula & Panhofer, 2018; Winnicott, 1960). Through these mechanisms, the presence of the group (Koch, Riege, et al., 2019), synchronization of the music, and creation of a specific group identity can facilitate these improvements. Unity of time, place, and space of the group also contributes to the development of participants’ sense of security (Sloan et al., 2013). Finally, combined with holding mechanisms, improvement in cultural support can be associated with individual development and cultural belonging intertwined with music and dance (Clarke et al., 2015). Cultural support and a sense of safety are even more important for people facing migration (Kira & Tummala-Narra, 2015). These key mechanisms should be highly regarded by clinicians and dance therapists, considering that the quality of the group is important in the resolution of psychological trauma (Koch, Riege, et al., 2019).

Finally, to improve the effectiveness of DT in the management of psychological trauma, clinicians, dance therapists, and facilitators should pay attention to specific settings inherent to the intervention. For example, the effects of physical activity through dance therapy depend on the context (White et al., 2017), the regularity, and the intensity of the activity (Chastin et al., 2021). Therefore, planning the dance therapy interventions provides a better therapeutic setting. Reaching a flow state also facilitates the management of psychological trauma as it reduces excessive self-consciousness (Riva et al., 2016). Within the group, professionals should be aware of their own skill set, and deploy interventions that encompass a better entrance in the flow state for their participants.

### Recommendations

Based on the results of this systematic review, some recommendation to research and practice can be considered. First, further quantitative, and qualitative research on the impact of dance therapy as a trauma-focused intervention should be conducted to complement the results of this qualitative systematic review. When Dance therapy is applied as a non-focused-trauma intervention, quantitative and qualitative research could further explore the emotional register, characteristics of the movement, and evolution of mental health afflictions associated with psychological trauma. Regarding DT practice for the management of psychological trauma, mental health professionals need to particularly ensure the stability of their intervention (unity of time, space, and place), the quality of the group, and their patients’ flow state.

### Strengths and limitations

The systematic article search was conducted on seven databases, in English and French. Quantitative, qualitative, and case studies were included, allowing for a fine-grained and complex analysis of the results. Bias analysis, performed by two researchers using two tools (NIH (National Institutes of Health, 2014) and JBI (Aromataris & Munn, 2020)), allowed the exclusion of studies with too much bias. The small number of available studies and the need for further quantitative research is the main limitation of our study.

## Conclusion

Dance therapy is an effective non-trauma-focused treatment, as it mobilizes the key dimensions for the management of psychological trauma (body, relaxation, diminution of stress, and interpersonal therapy). Its effectiveness is correlated with the therapist’s skill set, the group’s quality, the stability of the intervention, and the patient’s flow state. Conducting studies about the effect of DT as a trauma-focused intervention should be targeted in the future. Because of its setting, dance therapy appears particularly adapted to women survivors of violence, people facing migration, and clinical settings in which individuals have difficulty using language.

## Data Availability

All data produced in the present work are contained in the manuscript

## Acknowledgments and credits

We thank the LOBA association for granting us the funds and time necessary for this project.

## Author Biographies

**Crystal Tomaszewski** is a Ph.D. student at the University of Bourgogne/Franche-Comté and the University of Lorraine. Her research focuses on gender-based and sexual violence, psychological trauma, and dance-therapy interventions. She is interested in developing care devices to improve and diversify healthcare services, especially for women.

**Rose-Angélique Belot**, Ph.D., is an Associate Professor (HDR), in clinical psychology and psychopathology at the University of Bourgogne/Franche-Comté. Her research interests are vulnerability, body-psyche links, psychosomatic construction, trauma, and gender-based and sexual violence. She carries out numerous research projects in the field of health, in the Hospital’s neurology, maternity, and forensic medicine wards.

**Aziz Essadek**, Ph.D., is an Associate Professor in clinical psychology and psychopathology at the University of Lorraine. His research interests revolve around the consequences of child maltreatment and the specific psycho-therapeutic interventions for them.

**Héloïse Onumba-Bessonnet** is a co-facilitator in the RECREATION dance interventions of the LOBA association. She has been working for three years with migration-affected women victims of gender-based and sexual violence. She uses dance as a tool for the management of psychological trauma and comorbid afflictions.

**Christophe Clesse**, Ph.D., is a Clinical Psychologist and a Lecturer in Psychology at the University of Roehampton. His research interests focus on psychoneuroimmunology, women’s psychology, and body-psyche interactions as well as conducting research that aims at reducing the institutional, medical, and psychological inequalities that affect patients suffering from mental health issues.

